# Simulating the infected population and spread trend of 2019-nCov under different policy by EIR model

**DOI:** 10.1101/2020.02.10.20021519

**Authors:** Hao Xiong, Huili Yan

## Abstract

**Background:** Chinese government has taken strong measures in response to the epidemic of new coronavirus (2019-nCoV) from Jan.23, 2020. The number of confirmed infected individuals are still increasing rapidly. Estimating the accurate infected population and the future trend of epidemic spreading under control measures is significant and urgent. There have been reports external icon of spread from an infected patient with no symptoms to a close contact, which means the incubation individuals may has the possibility of infectiousness. However, the traditional transmission model, Susceptible-Exposed-Infectious-Recovered (SEIR) model, assumes that the exposed individual is being infected but without infectiousness. Thus, the estimating infected populations based on SEIR model from the existing literatures seems too far more than the official reported data.

**Methods:** Here, we inferred that the epidemic could be spread by exposed (incubation) individuals. Then, we provide a new Exposed-identified-Recovered (EIR) model, and simulated the epidemic spreading processes from free propagation phase to extremely control phase. Then, we estimate of the size of the epidemic and forecast the future development of the epidemics under strong prevention interventions. According to the spread characters of 2019-nCov, we construct a novel EIR compartment system dynamics model. This model integrates two phases of the epidemic spreading: before intervention and after intervention. We assume that 2019-nCov is firstly spread without intervention then the government started to take strong quarantine measures. Use the latest reported data from National Health Commission of the People’s Republic of China, we estimate the basic parameters of the model and the basic reproduction number of 2019-nCov. Then, based on this model, we simulate the future spread of the epidemics. Both the infected population and the spreading trend of 2019-nCov under different prevention policy scenarios are estimated. The epidemic spreading trends under different quarantine rate and action starting date of prevention policy are simulated and compared.

**Findings:** In our baseline scenario, the government has taken strict prevention actions, and the estimate numbers fit the official numbers very well. Simulation results tells that, if the prevention measures are relaxed or the action starting date of prevention measures is later than Jan. 23, 2020, the peak of identified individuals would be greatly increased, and the elimination date also would be delayed. We estimate the reproductive number for 2019-nCoV was 2.7. And simulation of the baseline scenario tells that, the peak infected individuals will be 49093 at Feb.16, 2020 and the epidemic spreading will be eliminated at the end of March 2020. The simulation results also tell that the quarantine rate and the starting date of intervention action policy have great effect on the epidemic spreading. Specifically, if the quarantine rate is reduced from 100% to less than 63%, which is the threshold of the quarantine rate to control the epidemic spreading, the epidemic spreading would never be eliminated out. And, if the starting date of intervention is delayed for 1 day than the date Jan. 23, the peak infected population will increase about 6351 individuals. If the delayed period is 3 days or 7 days, the increasing number would be 21621 or 65929 individuals, thus the peak infected number would up to 70714 and 115022 individuals.

**Interpretation:** Given that 2019-nCoV could be controlled under the strong prevention measures of what China has taken and it will take about three months. The confirmed infected individuals will still keep quick increasing for a generation period (27 days, equal to the sum of exposed period and identified period) after the start time point of control. The strong prevention measures should be insisted until the epidemics is wiped out. Other domestic places and overseas have confirmed infected individuals should take strong interventions immediately. Generally, earlier strong prevention measures could efficiently mitigate the outbreaks in other cities all over the world has confirmed individuals of epidemic of 2019-nCoV.

## 1 Introduction

In December 2019, atypical pneumonia caused by the zoonotic 2019 novel coronavirus (2019-nCoV) was first reported and confirmed in Wuhan, China. The novel coronavirus (2019-nCoV) is fast spreading, now to over 20 countries. In Jan.25, 2020, human-to-human transmission is confirmed[1-3].

To mitigate the spread of the virus, Chinese Government has implemented extremely serious quarantine prevention measures since Jan 23, 2020. All 31 provincial-level regions in China announced the highest public health alert. The extremely serious prevention measures are taken to mitigate the epidemic spreading, such as: Group tours, travel packages are suspended across China; Mass gatherings are all cancelled; Spring Festival holiday is extended; the start of Spring semester is postponed; Mask wearing is required at public place; temperature screening measures to detect individuals with fever have adopted at airports, train stations and Highway entrance, as well as entrance of community.

Until now, we still know little about the infectiousness of 2019-nCoV. The most worrisome aspect is the infectiousness of incubation (or exposed) individuals. That means, the incubation individual is being infected and maybe has the ability of infectiousness but without symptom. However, the existing forecasting model, Susceptible-Exposed-Identified-Recovered (SEIR) model, assumes that the exposed individual is being infected but without infectiousness. And other forecasting model, such as Susceptible-Infected-Recovered (SIR) model, even doesn’t consider the exposed process. So, maybe traditional transmission models of epidemic spreading are not suit for 2019-nCoV. That is a possible key reason for why the estimating infected population based on SEIR model are too far more than the official number. The estimate number is 75815 individuals in Jan. 25 2020 [4]while the official number is only 688 individuals; the estimate number is 190000 individuals in Wuhan in Feb. 4th 2020 [5] while the official number is only 1967 individuals. Furthermore, the forecasting model also should consider the effect of the prevention measures.

In this study, we provide a new Exposed-Identified-Recovered (EIR) model integrate the epidemic spreading processes before and after the prevention measures. And we supposed that the spreading process are mainly depended on the exposed process. On the contrary, as the identified individual generally will be quarantined by the hospital and can’t spread any more. Then, we nowcast the probable peak size of the epidemic and the possible duration of epidemic spreading, first by assuming the initial phase are freely spread by the exposed individuals, then the contact with the exposed individual are blocked under strict intervention measures that have been implemented from Jan.23, 2020. More importantly, from a public concern viewpoint, we simulate the possible future trend of the epidemic spreading under different quarantine rate and quarantine starting date.

## 2 Method

This article was intended to simulate the epidemic spreading trend under the first situation of free propagation and then under the intervention measures. We first constructed a new transmission model of epidemic spreading based on the system dynamics and SEIR model. Then we estimate the model parameters and the basic reproductive number of 2019-nCoV, on the basis of the latest official confirmed infected data in the mainland China. We then forecast the peak number of outbreak size and the duration of epidemic spreading under the extremely serious prevention measures. Finally, we simulate possible spreading course of 2019-nCoV under different quarantine rate and starting date of prevention measures in mainland China.

### Assumptions and model

Much is unknown about how 2019-nCoV spreads. Most often, spread from person-to-person happens among close contacts (about 6 feet). There have been reports external icon of spread from an infected patient with no symptoms to a close contact[6]. We inferred that the epidemic could be spread by exposed (incubation) individuals.

SEIR model has been widely used for modeling infectious diseases. However, it not suitable for the above spreading characters of the 2019-nCoV. We adjust this model and constructed EIR model. In which, the total population into three states: exposed (E, being infected as incubation with no symptom but with infectiousness), identified (I, being identified and quarantined without spread ability) and recovered (R, including death). In our model, exposed is assumed to be the first stage of infected individual, which is same as incubation stage, has no symptom and with infectiousness. So, we assume the exposed individuals has the possibility of infectiousness. And identified is assumed to be the second stage of infected individual, has symptom and be confirmed by hospital. So, we assume the identified individual has no ability of spreading since she/he will be pharmaceutical quarantined.

Furthermore, as differential equations are not suitable for the dynamic lag relationships between exposed individuals, identified individuals and recovered individual, the new recursive formulations are presented. The dynamical process of EIR thus could be described as:

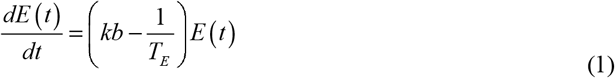

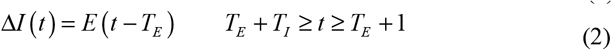

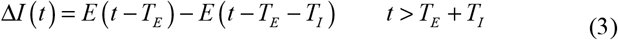

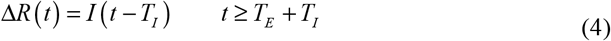

where *E*(*t*), *I*(*t*) and *R*(*t*) respectively represent the number of individuals in the exposed, identified and recovered states at date *t* (in days in later analyses). *T*_*E*_ and *T*_*I*_ were the mean exposed period (assumed to be the same as incubation) and treatment period; *b* is the risk of transmission per contact, *k* is the average contact by a person. Formulation (1) means a normal individual would turn to be an exposed individual with probability if she/he contacts with an infected individual; Formulation (2) and Formulation (3) means that, after each exposed period, an incubation individual would turn to be an identified individual. Formulation (4) means after each treatment period, an infected individual would be recovered or dead. The EIR epidemic spreading model is illustrated by figure 1.

**Figure 1:**
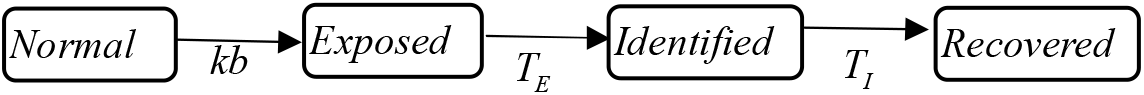
EIR epidemic spreading model.

From the dynamic formulations, we could easily deduce the formulations of the cumulated identified number, which are showed as formulation (5) and (6). The former is the first phase of cumulated before the generation period, which equal to the sum of exposed period and treatment period. The latter is the second phase of cumulated identified population after the generation period.

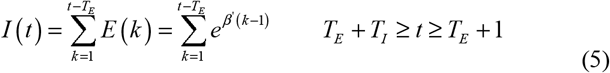

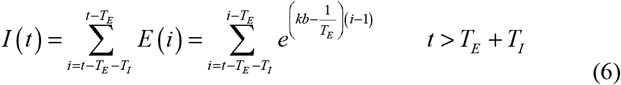

### Data sources

Early predictions for incubation time are between 2 and 14 days, based on data from similar coronaviruses, with the 95th percentile of the distribution at 12.5 days[1]. The 14-day criterion for epidemiological risk assumes the longest estimated incubation time[6]. So, we assume the exposed period is 14 days, so *T*_*E*_=14.

In the new publications of Li Qun et al. (2020) [1]and Huang et al. (2020)[3], the first case was confirmed on Dec. 1^st^ 2019. However, the official reported case is confirmed on Dec. 8, 2019 mentioned by Wuhan Municipal Health Commission[7]. In order to keep consensus with the previous researches, we prefer to take Dec 1^st^ 2019 as the date of the first infected individual identified. Thus, since the first exposed is identified on Dec. 1, 2019 after an exposed period (14 days), we inferred the first exposed individual is infected by the zoonotic source on Nov 17^th^ 2019 (*t*=1).

According to the official report data, the increased confirmed individuals are 259 in Jan 23th 2020 and the cured and discharged individuals are 261 in Feb 5^th^ 2020. Thus, we assume the infectious period is 13 days, *T*_*I*_=13.

From Jan 20^th^ 2020, the daily routine notification of infectious diseases is available on the website of National Health Commission of the People’s Republic of China. The main data is reported in Table 1.

**Table 1.** Official data of epidemic disease

| $t$ | date | Daily confirmed | Cumulated confirmed | Daily Cured | Daily Dead |
| --- | --- | --- | --- | --- | --- |
| 66 | Jan.20 | 77 | 291 | - | - |
| 67 | Jan.21 | 149 | 440 | - | 3 |
| 68 | Jan.22 | 131 | 571 | - | 8 |
| 69 | Jan.23 | 259 | 830 | 6 | 8 |
| 70 | Jan.24 | 444 | 1287 | 3 | 16 |
| 71 | Jan.25 | 688 | 1975 | 11(49) | 15(56) |
| 72 | Jan.26 | 769 | 2744 | 2 | 24 |
| 73 | Jan.27 | 1771 | 4515 | 9 | 26 |
| 74 | Jan.28 | 1459 | 5974 | 43 | 26 |
| 75 | Jan.29 | 1737 | 7711 | 21 | 38 |
| 76 | Jan.30 | 1982 | 9692 | 47 | 43 |
| 77 | Jan.31 | 2102 | 11791 | 72 | 46 |
| 78 | Feb.1 | 2590 | 14380 | 85 | 45 |
| 79 | Feb.2 | 2829 | 17205 | 147 | 57 |
| 80 | Feb.3 | 3235 | 20438 | 157 | 64 |
| 81 | Feb.4 | 3887 | 24324 | 262 | 65 |
| 82 | Feb.5 | 3694 | 28018 | 261 | 73 |
| 83 | Feb.6 | 3143 | 31161 | 387 | 73 |
| **84** | **Feb.7** | 3399 | **34546** | 510 | 86 |

### Parameter estimate

We assume the first infected incubation individual was on Nov. 17 2019, then Nov. 17 2019 is the first day of our simulations. So, *t*=1 corresponding to Nov. 17 2019. We assume average contact per person before strict prevention measures is 5 per day, that is *k*=5. From Jan. 23, 2020, corresponding to *t*=69, the extremely prevention measures have been taken, then the average contact person of exposed individuals are blocked, and has *k*=0. That is, when *t*>69, *k*=0.

According to the data source descripted above, we have *T*_*E*_=14, *T*_*I*_=13, *k*=5. Then, we can set parameter b for the EIR model to fit the most recent official cumulated data. Using the latest cumulated confirmed number 31161 Feb.6^th^ 2020 (*t*=83), we estimate the parameter *b*=0.038678. Hence, the basic reproduction number *R*_*0*_ *= k*b*T*_*E*_*=*2.7. And the transmission rate without prevention measures is β*=k*b= 0*.*1934*.

### Policy Projections

Most Chinese people were aware of the outbreak of 2019-nCoV by the mainstream media after 20 January 2020. The Hubei government released the announcement about strengthening the prevention and control measures against 2019-nCoV, and launched the second-level public health emergency response at 2:40am on 22 January, 2020. Thus, the public awareness and effective interventions were absent when the time was prior to this point. Then, Wuhan city limited inflow and outflow of people on January 23, 2020, which was the key measures to prevent and control the outbreak. From then on, more and more strong measures are taken to reduce the transmission of the epidemic.

As governments will take strict quarantine measures to prevent the pandemic of the new virus, we will model the policy results. We have different scenarios. Base Scenario: The government takes strict cases that all people who are found infected are strictly quarantined (k=0). Other Scenarios: The government takes strict cases that infected people are less strictly quarantined. These different scenarios corresponding to the ranges of *k* between 0 and 5 (see in table 2). *k*=0 indicates most strictly prevention measures are taken and quarantined rate is 100%. On the contrary, *k*= 5 means no prevention measure is taken and quarantined rate is 0%. Table 2 illustrates the relationship between quarantined rate and contact number *k*.

**Table 2.** Relationship between quarantined rate and the contact number *k*

| $k$ | 5 | 4.5 | 4 | 3.5 | 3 | 2.5 | 2 | 1.5 | 1 | 0.5 | 0 |
| --- | --- | --- | --- | --- | --- | --- | --- | --- | --- | --- | --- |
| Quarantined rate (%) | 0 | 10 | 20 | 30 | 40 | 50 | 60 | 70 | 80 | 90 | 100 |

## 3 Results

We expect that the epidemic of Corona virus in China will exist until May 2020. So, we set the simulation period is from Dec. 1, 2019 (*t*=1) to Apr. 30, 2020 (*t*=167). The baseline scenario is with quarantine rate 100% and action starting date Jan. 23, 2020 (*t*=69). Given different quarantine rates and action starting date, the main results are reported as follows.

Figure 2 showed the results of when quarantined rate is zero, that is *k*=5, which also could be taken as the scenario of no prevention measures are taken all the way. In this scenario, the number of the exposed individuals and identified individuals are presented as Exponential growth. At the end of April, there would be 740508835 individuals identified.

**Figure 2.**
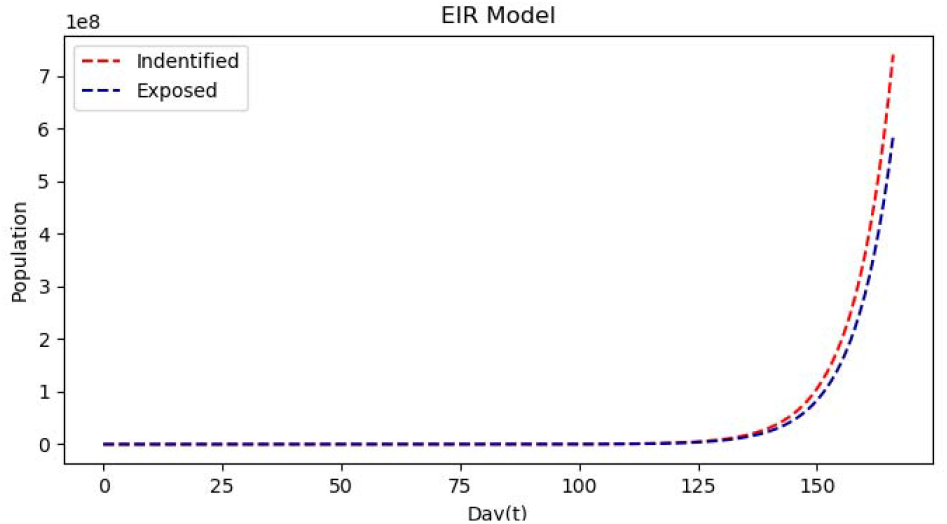
Epidemic spread trend without any control prevention measures.

Figure 3 showed the simulation curve of the baseline scenario from Dec. 1 2019 (*t*=1) to Apr. 30 2020 (*t*=167) and the trajectory of official data from Jan. 23, 2020 (*t*=66) to Feb. 7, 2020 (*t*=84). In figure 3, the green solid line is the trajectory of official data from Jan. 23, 2020 (*t*=66) to Feb. 7, 2020 (*t*=84), the red dashed line is the simulated curve of identified individuals. The second half of the trajectory of official data are well fitted by the simulation of the baseline scenario. From the simulation of baseline scenario, the estimation of the first half of the epidemic spreading from Jan. 20 (*t*=66) to Feb.7 (*t*=84) is higher than the official data. This could be explained by that, at the first stage, as the absence of the knowledge of the novel virus, the diagnoses are impossible to identify all the exposed individuals in time. So, it is naturally the confirmed number would be less than the actual exposed individuals at the beginning period of epidemic spreading.

**Figure 3.**
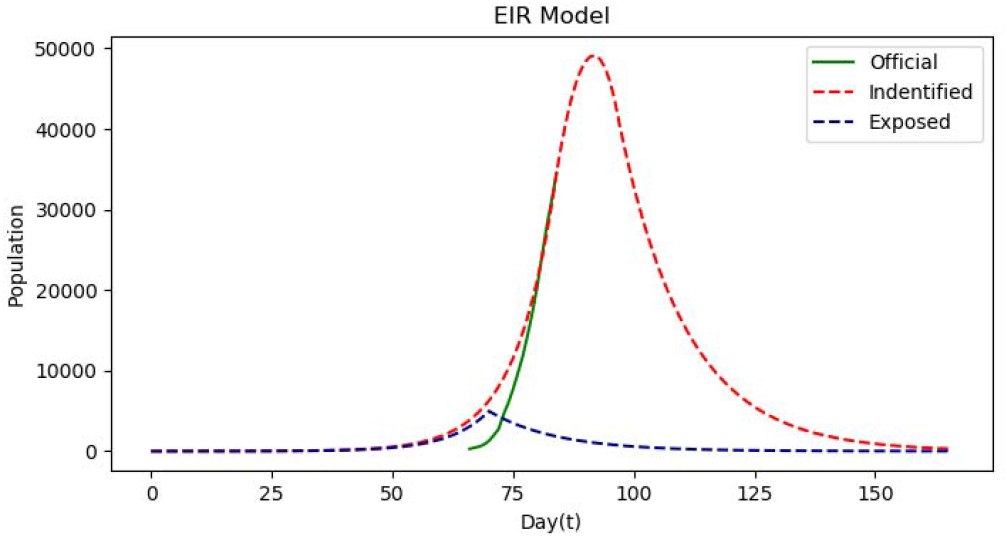
The estimate data and the official data from Jan 23 to Feb 7, 2020.

Figure 4 shows the epidemic trend of the cumulated exposed number and identified number with average contact number *k* reduced from 5 to 0, 0.5, 1, 1.5, 1.852 and 2, which corresponding to quarantined rate 100%, 90%, 80%, 70%, 63%, 60%, after control quarantined measures have been taken from Jan 23, 2020 (*t*=69, not include 69). Table 3 shows the peak confirmed population and the date. We estimated that, if there are the extremely quarantined measures from Jan 23th 2020, the peak confirmed infected population will be 49093 on Feb.16, 2020. And the epidemics would be eliminated in the end of April. If the prevention measures are relaxed and the quarantined rate is reduced, then the peak number would be substantially increased and the peak date also would be delayed. The epidemics would be controlled and wiped out if average contact number *k* was reduced less than 1.85, the quarantined rate more than 63%.

**Table 3.** Estimated results under different quarantined rate

| Quarantined rate | Peak date | Peak confirmed population |
| --- | --- | --- |
| 100% | Feb.16 | 49093 |
| 90% | Feb.17 | 51605 |
| 80% | Feb.18 | 55059 |
| 70% | Feb.19 | 59953 |
| 63% | Feb.20 | 64740 |

**Figure 4.**
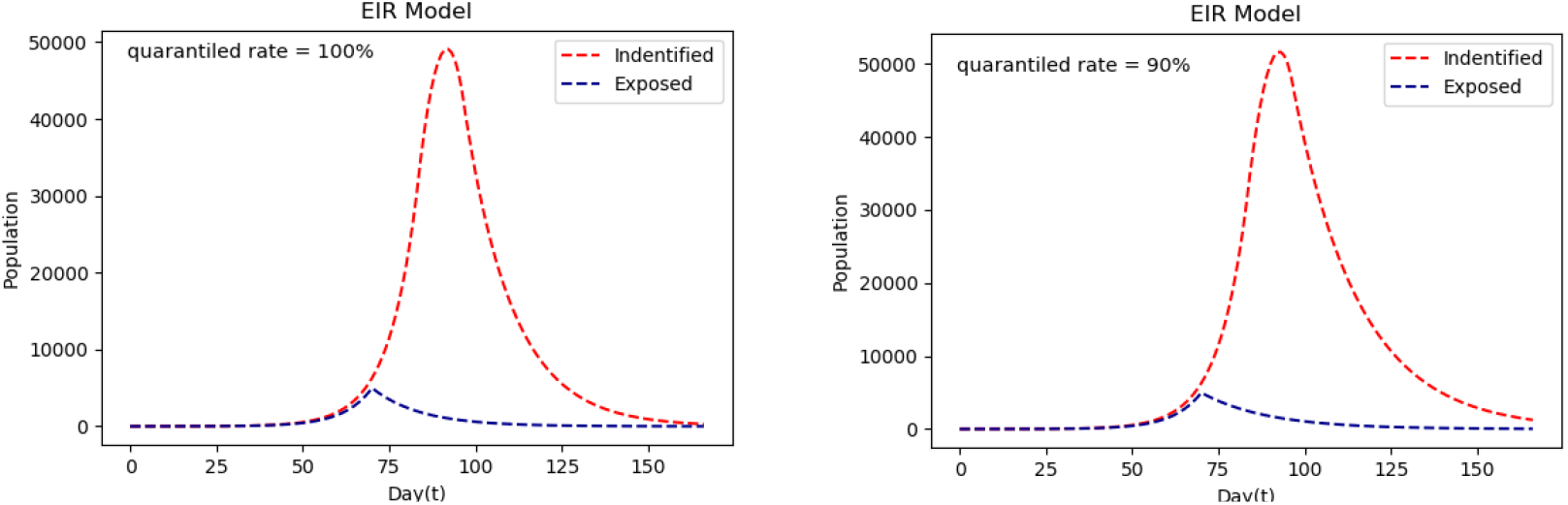
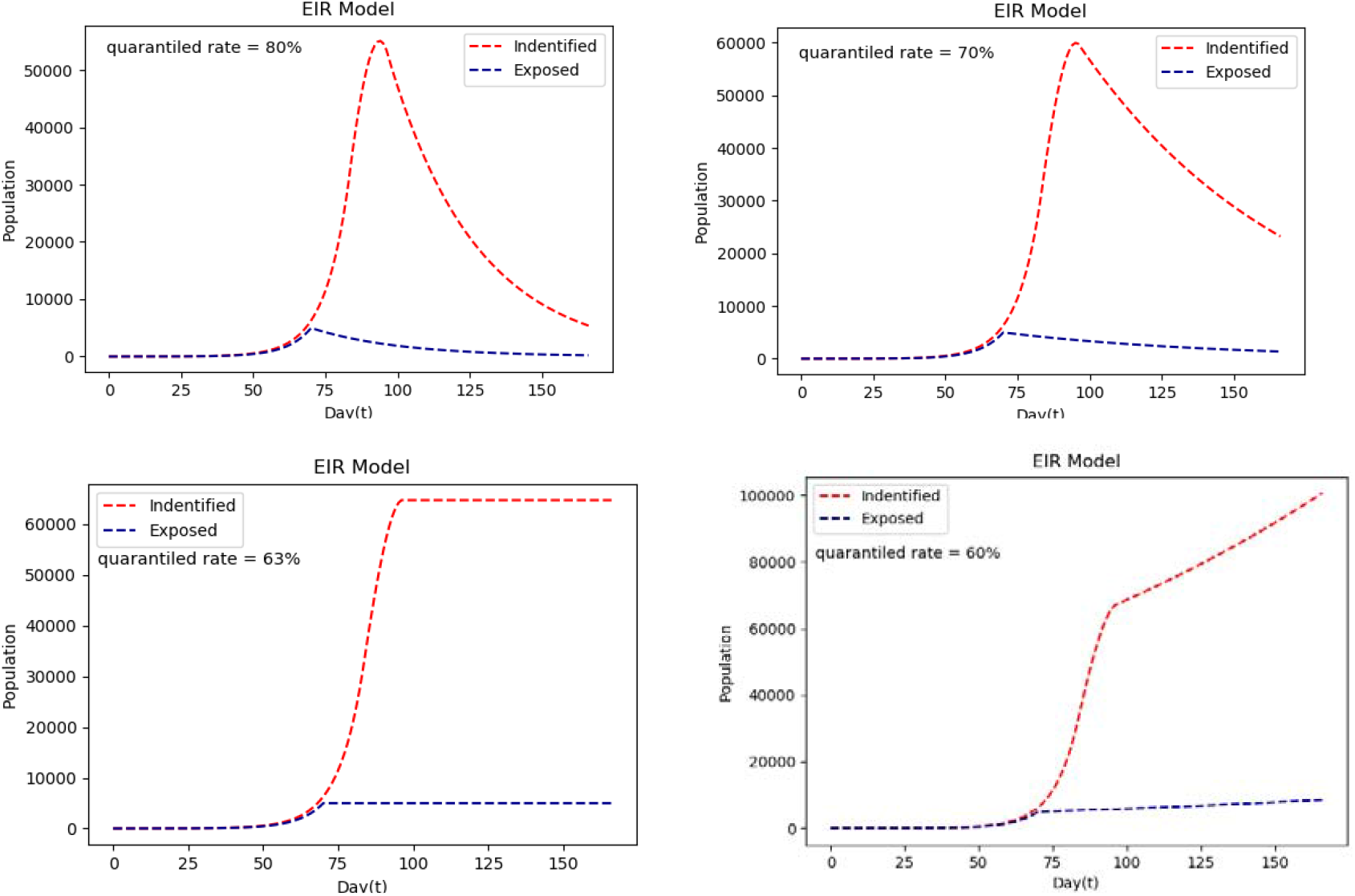
Epidemic spread trend in different policy scenarios.

Compared to the action start date Jan. 23rd 2020, if the action starting date is taken earlier or delayed 1 day, 3 days or 7 days, figure 5 and table 4 shows the simulation results. Compare to the baseline action date, Jan. 23, 2022, one day ahead would reduce 5622 identified individuals and one day delay would increase 6351 identified individuals. If the action starting days are delayed for 3 days or 7 days, the increasing identified individuals would up to 21621 and 65929, which means the peak number of identified individuals would be up to 70714 and 115022.

**Table 4.** Estimated results under different quarantined date

| Prevention date | Peak confirmed population | Peak date |
| --- | --- | --- |
| Jan.16 | 20953 | Feb.9 |
| Jan.20 | 34083 | Feb.13 |
| Jan.22 | 43471 | Feb.14 |
| **Jan.23** | **49093** | **Feb.16** |
| Jan.24 | 55444 | Feb.17 |
| Jan.26 | 70714 | Feb.19 |
| Jan.30 | 115022 | Feb.23 |

**Figure 5.**
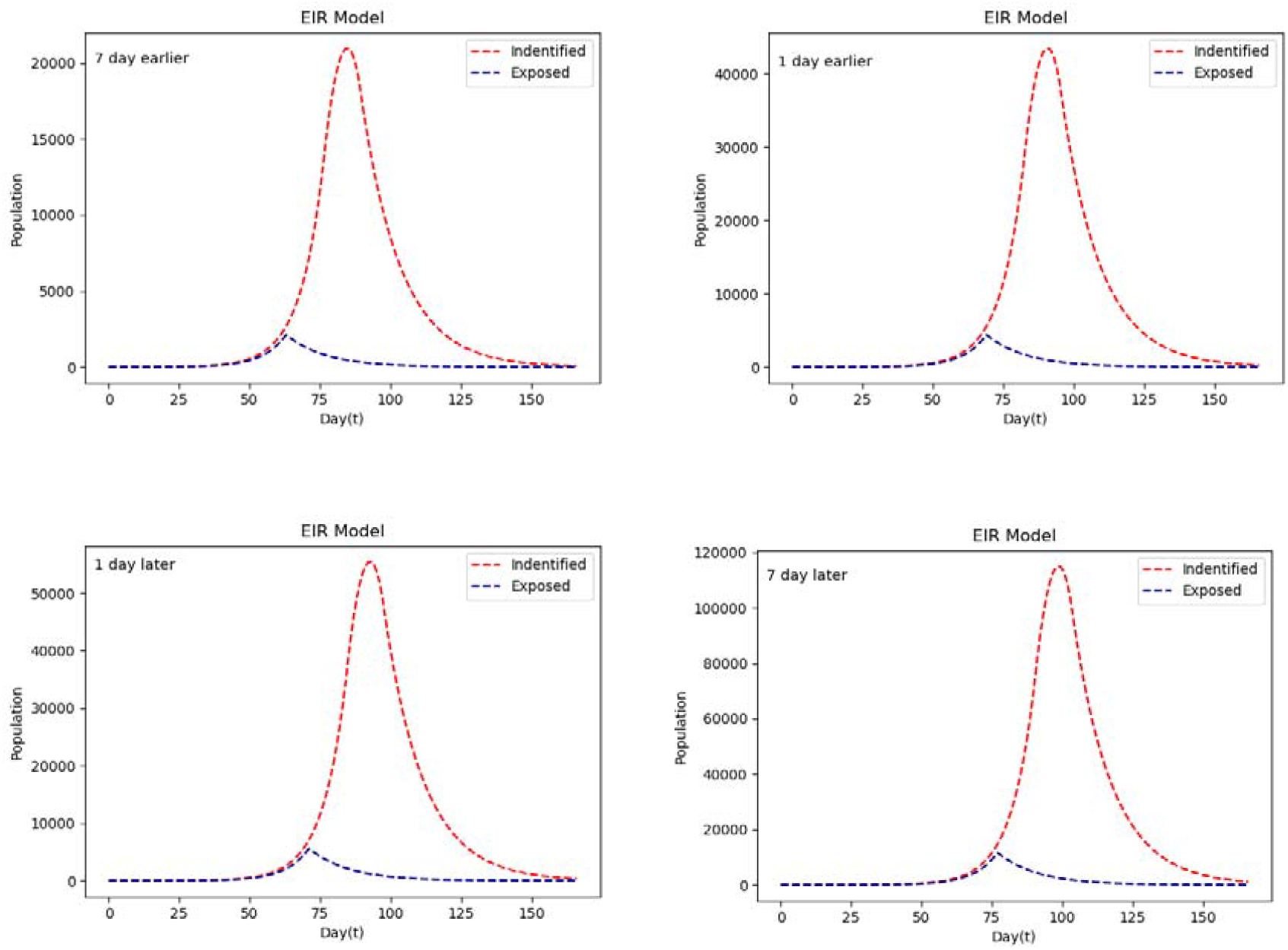
Epidemic spread trend in different policy scenarios.

## Discussion

Since Jan 23rd 2020, after outbreak of the novel atypical pneumonia, government has applied substantial draconian intervention measures to drastically reduce within-population contact rates. These measures include extending the Spring Festival holiday, and instituting work-from-home arrangements, postpone the start of Spring semester, Nurseries and early learning centers closures, cancellation of mass gatherings, wearing masks at public palce, and so on. However, the reported case numbers are still rising rapidly. Simulating the epidemic spreading trend of 2019-nCoV under the control measures are of crucial importance for public health planning and control domestically and internationally. Several earlier publications have given some useful forecasting of 2019-nCoV [4, 5, 8-11], but their estimating numbers are too far more than the official data. The estimating of epidemic spreading should consider the effect of actions have been taken to mitigate the spreading. Furthermore, the asymptomatic infection and transmission characters should be considered in the transmission model. Transmission model and parameters are essential for the generation of accurate forecasting of infected population and spreading trend [12].

In this study, we have constructed a novel EIR transmission model to stimulate the epidemic spreading. On the trajectory of simulating curve of baseline scenario, the peak number of infected population of 2019-nCoV will be 49093 on Feb 16, and it could be securely contained the spread of infection under the current draconian intervention measures. However, these intervention measures should be kept for about 2 months until the end of April. And extremely draconian level prevention measures should be kept at least an incubation period after the infected population peak. In the baseline scenario, the government takes the strict prevention actions, the estimate numbers fit the official numbers very well. There can be no doubt that the official numbers are accurate.

Simulating result also tells that the quarantined rate, which present the degree of the prevention measures, has great effect on the peak number and duration of the epidemic spreading. Only the quarantined rate is higher than 80%, the epidemic spreading could be reduced drastically and the identified population would be less than 5000 in the end of March 2020. Even at the threshold degree, when the quarantined rate is 63%, the infected population would up to a peak number 115022 and keep at that number for ever unless more strict prevention measures are taken. If the quarantined rate is less than 63% the epidemic spreading of 2019-nCoV will never be eliminated and the infected population would still keep increasing.

The simulation results of different action starting date support the common sense that the control action should be taken as early as possible. Specifically, simulation results tells that the action starting date of prevention measures later than Jan. 23, 2020, the peak of identified individuals would be greatly increased. And the wipe out date also would be delayed.

The modelling techniques that we used in this study are based on SEIR and system dynamics. And our model is parameterized with the latest official released data of 2019-nCoV. An additional strength of our study is that simulation of the baseline scenario is well fit the trajectory of the official data of epidemic spreading. Nonetheless, our study has several major limitations. First, we assumed that the transmission of 2019-nCoV mainly caused by exposed individuals (without symptoms) and the transmission ability of identified (without symptoms) is ignored. Second, our transmission model was somewhat sensitive to several key parameters: the starting date of epidemic spreading, the exposed (incubation) period and the identified (treatment) period. And our assumption regarding the exposed (incubation) period and the identified (treatment) period is keeping stable. If 2019-nCoV, similar to influenza, has strong seasonality in its transmission, our epidemic forecast might not be reliable.

## Contributors

Huili Yan designed the experiments and collected data. Hao Xiong analysed data, interpreted the results and wrote the manuscript.

## Data Availability

All the original data are included manuscript. And the simulating data could be available if needed

## Data sharing

Data obtained for this study will not be made available to others.

## Acknowledgments

This work was supported by National Natural Science Foundation of China (Grant No. 71761009, No. 71461007 and No. 71461006) and Hainan Province Planning Program of Philosophy and Social Science (HNSK(YB)19-06, HNSK(YB)19-11).

